# Non-invasive Auricular Vagus nerve stimulation for Subarachnoid Hemorrhage (NAVSaH): Protocol for a prospective, triple-blinded, randomized controlled trial

**DOI:** 10.1101/2024.03.18.24304239

**Authors:** Anna L Huguenard, Gansheng Tan, Gabrielle W Johnson, Markus Adamek, Andrew T Coxon, Terrance T Kummer, Joshua W Osbun, Ananth K Vellimana, David D. Limbrick, Gregory J Zipfel, Peter Brunner, Eric C Leuthardt

## Abstract

**Background:** Inflammation has been implicated in driving the morbidity associated with subarachnoid hemorrhage (SAH). Despite understanding the important role of inflammation in morbidity following SAH, there is no current effective way to modulate this deleterious response. There is a critical need for a novel approach to immunomodulation that can be safely, rapidly, and effectively deployed in SAH patients. Vagus nerve stimulation (VNS) provides a non-pharmacologic approach to immunomodulation, with prior studies demonstrating VNS can reduce systemic inflammatory markers, and VNS has had early success treating inflammatory conditions such as arthritis, sepsis, and inflammatory bowel diseases. The aim of the Non-invasive Auricular Vagus nerve stimulation for Subarachnoid Hemorrhage (NAVSaH) trial is to translate the use of non-invasive transcutaneous auricular VNS (taVNS) to spontaneous SAH, with our central hypothesis being that implementing taVNS in the acute period following spontaneous SAH attenuates the expected inflammatory response to hemorrhage and curtails morbidity associated with inflammatory-mediated clinical endpoints.

**Materials and methods:** The overall objectives for the NAHSaH trial are to 1) Define the impact that taVNS has on SAH-induced inflammatory markers in the plasma and cerebrospinal fluid (CSF), 2) Determine whether taVNS following SAH reduces radiographic vasospasm, and 3) Determine whether taVNS following SAH reduces chronic hydrocephalus. Following presentation to a single enrollment site, enrolled SAH patients are randomly assigned twice daily treatment with either taVNS or sham stimulation for the duration of their intensive care unit stay. Blood and CSF are drawn before initiation of treatment sessions, and then every three days during a patient’s hospital stay. Primary endpoints include change in the inflammatory cytokine TNF-α in plasma and cerebrospinal fluid between day 1 and day 13, rate of radiographic vasospasm, and rate of requirement for long-term CSF diversion via a ventricular shunt. Secondary outcomes include exploratory analyses of a panel of additional cytokines, number and type of hospitalized acquired infections, duration of external ventricular drain in days, interventions required for vasospasm, continuous physiology data before, during, and after treatment sessions, hospital length of stay, intensive care unit length of stay, and modified Rankin Scale score (mRS) at admission, discharge, and each at follow-up appointment for up to two years following SAH.

**Discussion:** Inflammation plays a central role in morbidity following SAH. This NAVSaH trial is innovative because it diverges from the pharmacologic status quo by harnessing a novel non-invasive neuromodulatory approach and its known anti-inflammatory effects to alter the pathophysiology of SAH. The investigation of a new, effective, and rapidly deployable intervention in SAH offers a new route to improve outcomes following SAH.

**Trial registration:** Clinical Trials Registered, NCT04557618. Registered on September 21, 2020, and the first patient was enrolled on January 4, 2021

## Introduction

### Background and rationale

Intracranial aneurysms are common, with 3-5% of all adults harboring at least one. The resulting subarachnoid hemorrhage (SAH) from ruptured aneurysms accounts for 5–10% of all strokes worldwide, culminating in a total of 600,000 new cases per year [1]. SAH is a major driver of mortality and morbidity, with 10-25% of patients dying following SAH and an additional 30% of patients suffering permanent disability [2]. While the immediate sequelae of SAH can include risk for re-rupture, elevated intracranial pressure, and acute hydrocephalus, secondary injury is a major driver of morbidity as mediated by early brain injury, cerebral vasospasm, delayed cortical ischemia, and chronic hydrocephalus [3]. Targeting the post-hemorrhage period with the goal of reducing these secondary sequelae from SAH is an important mechanism for improving outcomes in SAH patients.

There is growing evidence that systemic and local inflammation may promote aneurysm formation and rupture as well as lead to poorer outcomes following aneurysmal rupture. Following SAH, blood within the subarachnoid space triggers both local, as measured by inflammatory markers in the cerebrospinal fluid (CSF), and systemic inflammatory responses, as measured by inflammatory markers in the blood. Prior work has identified increased cytokines including IL-1β, IL-4, IL-8, IL-10, IL-18, and IL-33, following SAH in animal [4–7] and human [8–13] studies. Key drivers of SAH-induced inflammation are the cytokines IL-6 and TNF-α. Studies in canine and rabbit models of SAH demonstrate increased rates of vasospasm correlated with expression of IL-6 in the basilar artery [6] and CSF [14], respectively. Work performed in humans reveals elevated IL-6 in the blood [11] and CSF [9,11,15-17] following SAH. In several studies, elevated IL-6 in the CSF was significantly correlated with development of vasospasm [9,16], delayed cortical ischemia [17], chronic hydrocephalus [18,19], and poorer overall outcome [9,11] following SAH. Similarly, studies in rodent [20] and rabbit [21] models of SAH found increased TNF-α was associated with vasospasm. In humans, elevated TNF-α in the blood [22,23] and CSF [8,9,24,25] have been demonstrated following SAH. Elevated TNF-α has been associated with vasospasm [8,9,24], hydrocephalus [8], and poor outcome [8,22].

Numerous anti-inflammatory interventions have been trialed in humans to better understand, and eventually target, inflammatory pathways to improve outcomes after SAH [26–29]. In smaller enrollment studies, there was promise of outcome improvement with Cyclosporine A [30,31] and steroids like methylprednisolone [32,33], hydrocortisone [34], and dexamethasone [35]. Other medications demonstrated no impact on overall outcomes, like Clazosentan [36,37], Cilostazol [38], and IL-1 antagonists [39,40]. In several meta-analysis studies looking at groups larger than 1000 patients, Simvastatin [41], Aspirin, non-steroidal anti-inflammatory medications, and thienopyrindines [42] all demonstrated no improvement in outcomes. In summary, while some pathway-targeted pharmacological approaches have led to changes in secondary outcomes of vasospasm and delayed cerebral ischemia in clinical trials [37,38], these approaches have failed to produce an effective intervention that reliably improves outcomes in SAH patients. Thus, there is an urgent need to find a novel approach to more globally and meaningfully reduce inflammation in SAH to reduce patient morbidity.

Vagus nerve stimulation (VNS) has been studied as a novel method of reducing inflammation. Substantial work has demonstrated that products of infection or injury activate sensory neurons traveling to the brainstem in the vagus nerve [43,44]. The arrival of these incoming signals generates action potentials that travel from the brainstem to the spleen and other organs. This culminates in T cell release of acetylcholine, which interacts with α7 nicotinic acetylcholine receptors (α7 nAChR) on immunocompetent cells to inhibit cytokine release in macrophages [45]. This neural-immunomodulatory circuit, referred to as the “cholinergic anti-inflammatory pathway”, presents opportunities for developing novel therapeutic strategies to treat inflammatory diseases. It has been successfully implemented in models of inflammatory conditions like induced neuroinflammation [46], cerebral ischemia/reperfusion [47], rheumatoid arthritis [48], sepsis [49], and inflammatory bowel diseases [50,51] or colitis [52]. Harnessing its anti-inflammatory effects, VNS has been used in a mouse model of cerebral aneurysms and SAH [53]. In this study, pre-treatment with VNS not only reduced the rupture rate of intracranial aneurysms, but also reduced the grade of hemorrhage if rupture occurred and improved survival and outcome after SAH [53]. There has not been any published work implementing VNS following SAH in humans.

Historically, VNS was performed exclusively by surgical cervical neck dissection and placement of a cuff electrode directly around the nerve within the carotid sheath. Alternatively, VNS can be accomplished non-invasively by stimulating the auricular branch of the vagus nerve as it courses through the external ear, obviating the morbidity of a surgical procedure and allowing rapid deployment of the intervention in critically ill patients. The external ear is an ideal target for non-invasive stimulation of the vagus nerve, where the auricular branch travels in the concha of the ear [54]. This transcutaneous auricular approach has demonstrated good efficacy, with minimal morbidity [54,55].

Taken together, there is substantial evidence that VNS is an emerging tool to mitigate inflammation, but there is a dearth of understanding on its effects on SAH in humans.

### Objectives

Our long-term goal is to translate the use of non-invasive transcutaneous auricular VNS (taVNS) to reduce morbidity and improve outcomes in patients following spontaneous SAH. The overall objectives for the NAVSaH trial in pursuit of achieving this goal are to (i) demonstrate the impact taVNS has on inflammatory markers in the blood and CSF in patients following SAH, and (ii) determine if taVNS reduces the incidence of inflammation-mediated sequelae of SAH by performing a prospective, randomized controlled trial. Our central hypothesis is that implementing taVNS in the acute period following spontaneous SAH will attenuate the expected inflammatory response to hemorrhage and will curtail morbidity associated with inflammatory-mediated clinical endpoints (i.e., vasospasm & hydrocephalus). By pursuing this project, we expect to demonstrate the important role that neuromodulation can play in SAH. More specifically, we expect this work will create the foundation of knowledge for advancing non-invasive taVNS to enable further research to translate this as an established clinical intervention in the future.

## Materials and methods

### Trial design

NAVSaH is a prospective, triple-blinded (patient, care team, and outcomes assessor), randomized-control trial to assess superiority of intervention with vagus nerve stimulation following spontaneous subarachnoid hemorrhage compared to standard of care with respect to inflammatory markers in the blood and CSF, rate of radiographic vasospasm, and rate of development of chronic hydrocephalus. Treatment and sham assignments are made with a 1:1 allocation. Figs 1 and 2 represents an overview of the trial design. This protocol manuscript was written in accordance with the Standard Protocol Items: Recommendations for Interventional Trials (SPIRIT) guidelines [56] and the Consolidated Standards of Reporting Trials (CONSORT) statement [57].

**Fig 1:**
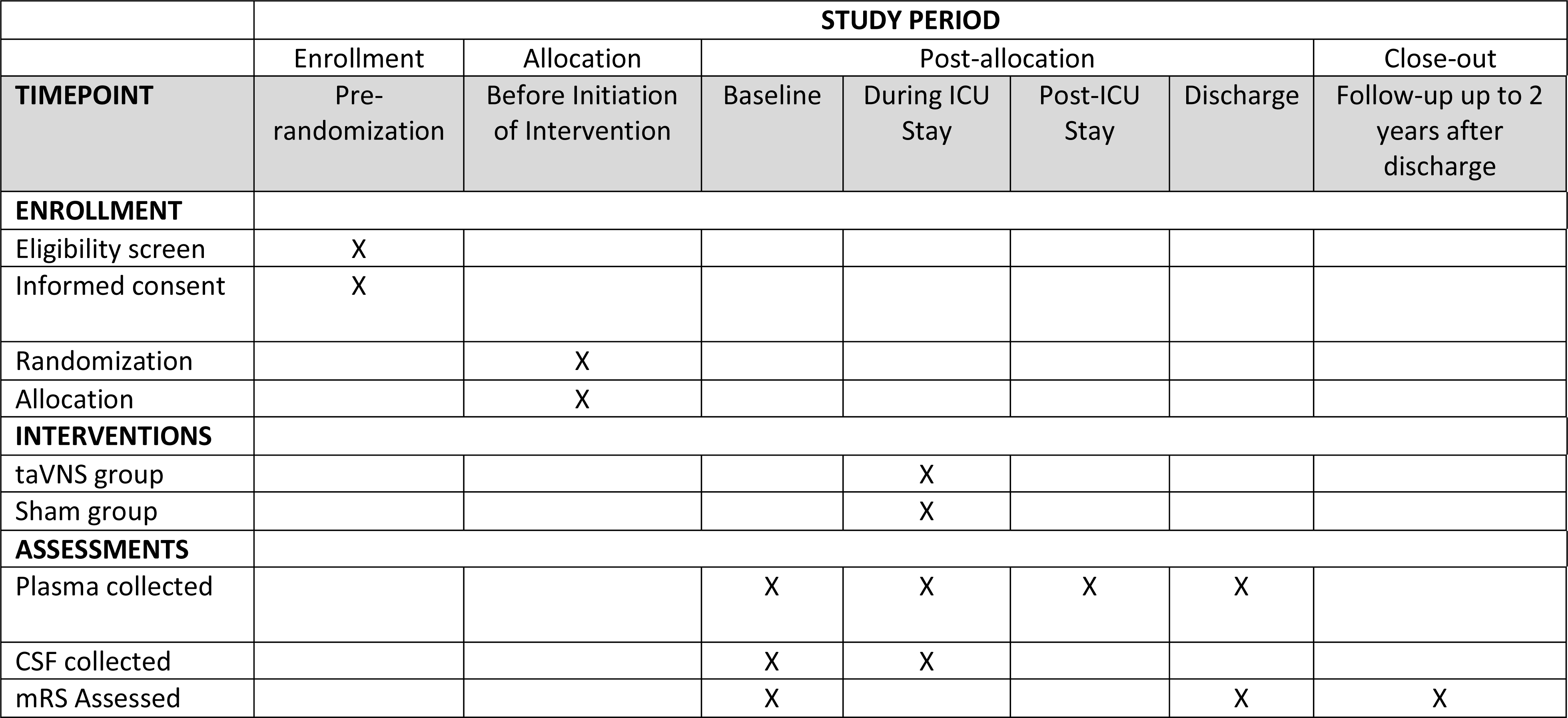
SPIRIT schedule of enrollment, interventions, and assessments.

**Fig 2:**
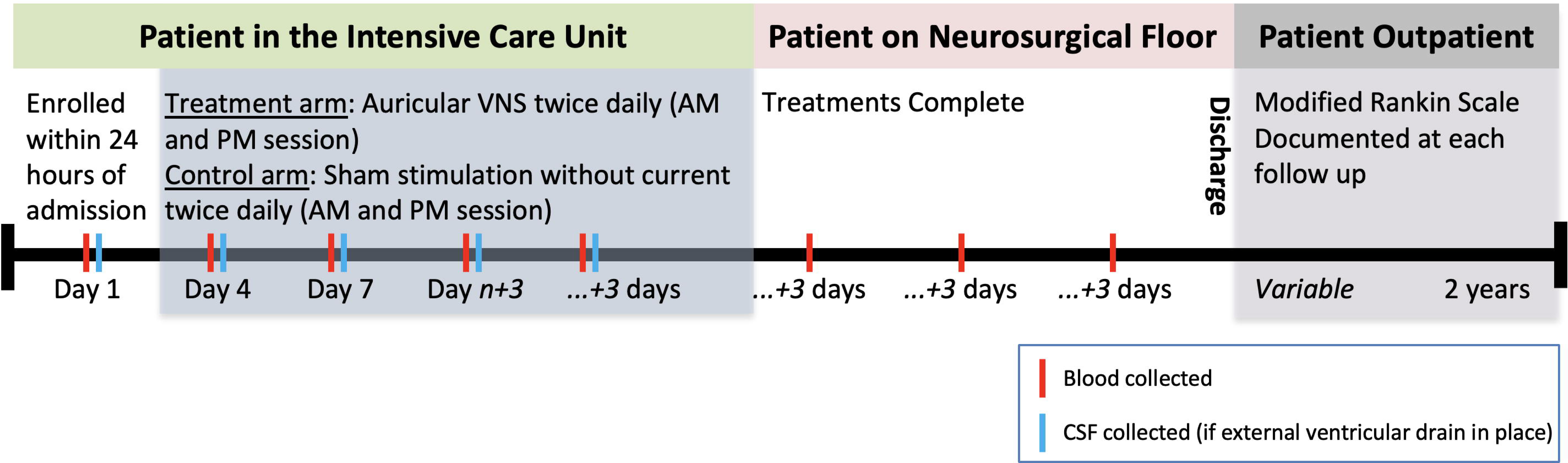
Timeline for enrollment and participation in the NAVSaH clinical trial by participants. Cerebrospinal fluid (CSF)

## Methods: participants, interventions and outcomes

### Study setting

The NAVSaH trial is being conducted at a single center, a tertiary care academic hospital center, Barnes Jewish Hospital/Washington University in St. Louis (St. Louis, MO, USA).

### Eligibility criteria

Participants include all individuals who present with a spontaneous subarachnoid hemorrhage, aged >18 years old. Exclusion criteria includes traumatic etiology for hemorrhage, negative vascular imaging for aneurysm (CT angiogram or cerebral angiogram), ongoing cancer therapy, ongoing use of an immunomodulating or suppressive medication, sustained bradycardia on arrival with a heart rate < 50 beats per minute for > 5 minutes, implanted pacemaker or other electrical device, current pregnancy, or a positive test for the Covid-19 virus.

### Who will take informed consent?

Patients provide written consent for themselves when able. For patients who are unable to provide consent given cognitive impairment from their subarachnoid hemorrhage, then a legally authorized representative can provide written informed consent in conjunction with patient assent (whenever possible). The patient or legally authorized representative are informed about the objectives of the study as well as potential risks and benefits of participation. Documentation of consent is kept in paper format and maintained in a locked and secure storage box by trial staff. Consent is obtained by either a physician member of the research team, or an appropriately trained research coordinator. In all situations, the team member seeking informed consent is not involved in the medical care of the enrollee, to prevent actual or implied bias or compulsion for trial participation.

### Additional consent provisions for collection and use of participant data and biological specimens

There are no additional consent provisions for specific ancillary studies, however, there is an optional provision in the consent form to allow for use of data collected for future related studies.

### Interventions

#### Explanation for the choice of comparators

Both arms of the NAVSaH trial will receive standard of care for management of subarachnoid hemorrhage, not limited to intensive care monitoring, treatment of a ruptured aneurysm via endovascular or surgical means as dictated by the physician team, cerebrospinal fluid diversion when considered medically indicated, and repeated imaging studies as deemed necessary by the physician team. Treatment with non-invasive vagus nerve stimulation has been implemented in many other studies, and has a demonstrated safety profile when implemented previously following acute ischemic or hemorrhagic stroke [58]. Sham stimulation, which is delivered as a comparator, involves the placement of ear clips without electrical stimulation has no perceived positive or negative effect on clinical outcome, or foreseeable risk for adverse effects.

#### Intervention description

Enrolled participants are randomized to one of two study arms and receive either 1) twice daily non-invasive auricular vagus nerve stimulation to the left ear, or 2) sham stimulation involving placement of the ear clips without applied electrical current. Treatment is applied twice daily, similar to other studies applying vagus nerve stimulation to drive reduction in inflammation [59–61]. The electrodes are placed on the left ear, consistent with the typical left-sided cervical placement of invasive vagus nerve stimulators [62], although no evidence indicates laterality in cardiac effects in non-invasive stimulation. An enrolled patient will begin with either taVNS or sham stimulation twice daily during their stay in the intensive care unit while continuous heart rate/rhythm, blood pressure, intracranial pressure, and oxygenation monitoring is in place. Each day, there is one session each morning (between 05:00-10:00) and one each evening (between 16:00-21:00).

The initial treatment session occurs during the first treatment block time after informed consent is complete. All patients are fitted with a portable TENS (transcutaneous electrical nerve stimulation) unit connected to two ear clips, applied to the left ear during treatment periods. For VNS treatment, these ear clips are placed along the concha of the ear (Fig 3), with positioning optimized to target the auricular branch of the vagus nerve [61]. Stimulation parameters were selected based on prior studies that sought to maximize vagus somatosensory evoked potentials while avoiding perception of pain [63,64]. Stimulation parameters will be 20 minutes duration, frequency of 20 Hz, 250μs pulse width, and an intensity of 0.4mA. Sham treatments will involve no electrical current. Treatment sessions with either taVNS or Sham stimulation will end after transfer out of the intensive care unit.

**Fig 3:**
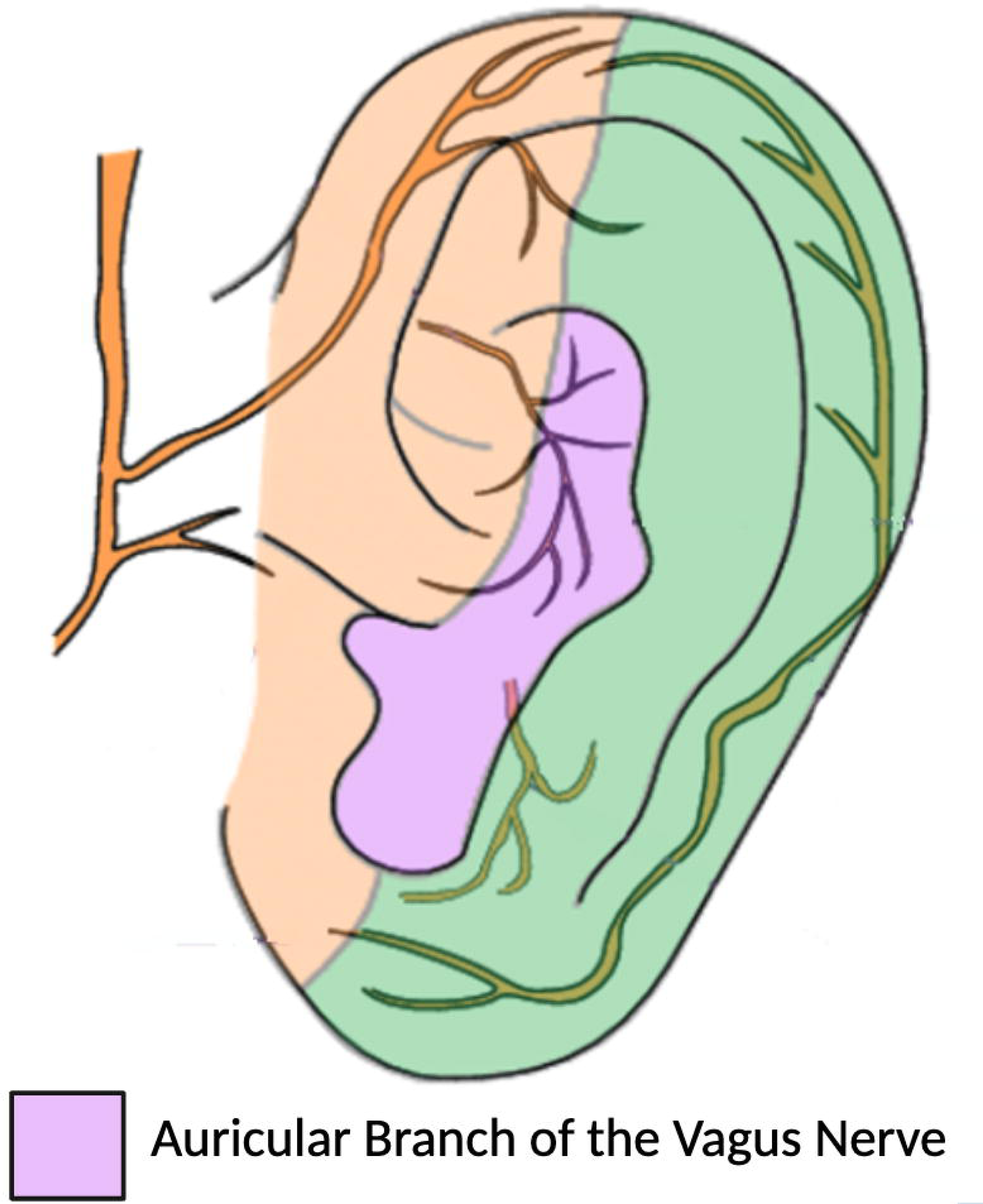
Target for auricular vagus nerve stimulation. Vagus nerve stimulation is administered by placing the stimulating electrodes along the concha of the ear, where the innervation of the auricular branch of the vagus nerve runs.

#### Criteria for discontinuing or modifying allocated interventions

The study treatment will be discontinued if a participant meets any of the following criteria: if irritation or redness at the stimulation site occurs, evidence of induced clinically significant changes in heart rate or rhythm, or by participant or legally authorized representative request. In these cases, the treatment sessions will cease, but blood and cerebrospinal fluid draws and recording of clinical outcomes will continue unless by request by the patient or legally authorized representative request.

#### Strategies to improve adherence to interventions

All treatment sessions occur while the enrolled patient is admitted to the hospital, and in the intensive care unit. Therefore, we anticipate few barriers to adherence with the intervention.

#### Relevant concomitant care permitted or prohibited during the trial

All medications, procedures, or interventions recommended by the medical team to manage an enrolled patient’s subarachnoid hemorrhage will be permitted. The patient will not be permitted to receive any intervention that is associated with a separate clinical trial.

#### Provisions for post-trial care

There is no anticipated harm and no planned compensation for trial participation. All post-trial care will follow the typical follow-up for patients discharged after subarachnoid hemorrhage (including follow-up imaging or physician office visits). No additional imaging, laboratory studies, or hospital visits are otherwise required due to trial participation alone.

### Outcomes

The NAVSaH trial’s evaluated end points include: 1) Defining the impact that taVNS has on the SAH-induced inflammatory markers in the plasma and cerebrospinal fluid (CSF), 2) Determining if taVNS following SAH reduces angiographic vasospasm 3) Determining if taVNS following SAH reduces chronic hydrocephalus.

For assessment of the impact taVNS has on inflammatory makers, blood and CSF is collected prior to the initial treatment session as a baseline, and then every three days during the patient’s inpatient hospital stay (CSF collected only when an external ventricular drain is in place). The primary inflammatory marker assessed is TNF-α, although a panel of 13 cytokines is analyzed in an exploratory fashion (IL-1β, IL-2, IL-4, IL-5, IL-6, IL-8, IL-10, IL-12, IL-13, IL-17, TNF-α, GM-CSF, and IFN-γ). Differences regarding the local and systemic inflammatory cytokine TNF-α reduction from baseline to day 13 post-treatment will primarily be examined through the interaction of time and treatment effects, with additional exploratory analyses performed for other days.

To assess angiographic vasospasm, serial vascular imaging studies will be evaluated. In addition to initial diagnostic imaging, patients will undergo a repeat computed tomography or catheter angiogram seven days after admission, per our hospital’s standard protocol. Additionally, further vascular imaging will be performed if there is clinical concern per the intensive care or neurosurgical teams for clinical vasospasm or stroke. For both planned and indicated imaging sessions, each vascular imaging study will be reviewed by a trained neurointerventionalist blinded to treatment arm, who will measure and quantify the imaging as it relates to vasospasm as none, mild (< 25% stenosis), moderate (25%–50% stenosis), or severe (> 50% stenosis) narrowing of at least one major intracranial artery, as previously described [65,66].

To evaluate the rate of chronic hydrocephalus, the primary outcome assessed will be binary (placement of ventricular shunt, versus no shunt placement).

The trial’s secondary end points include: patient demographics (gender, ethnicity, and age), number and type of hospitalized acquired infections, duration of external ventricular drain in days, interventions required for vasospasm (blood pressure augmentation, intraarterial medications, intrathecal medications, endovascular intervention with angioplasty), continuous physiology data (heart rate, blood pressure, respiratory rate, intracranial pressure) before, during, and after treatment sessions, hospital length of stay, intensive care unit length of stay, and modified Rankin Scale score (mRS) at admission, discharge, and each at follow-up appointment for up to two years following SAH.

### Participant timeline

The schematic overview of the NAVSaH trial is shown in Fig 2. Study start-up began in July 2020, including protocol development and single institutional review board approval. Trial enrollment began in January 2021, and it is anticipated that enrollment will continue for approximately 4 years from that date. Participants are enrolled in an acute care setting when they present with a spontaneous SAH, with enrollment occurring within 24 hours of arrival to the hospital. Intervention with taVNS or Sham stimulation occurs twice daily while the patient is in the intensive care unit, and discontinues on transfer to lower level of care from ICU. A mRS is assessed at time of hospital admission, at time of discharge, and at all follow-up appointments with the Neurosurgery clinic or Neuroangiography team in the two years following admission by an assessor blinded to treatment arm. Follow-up with the Neurosurgical and Neuroangiography teams follows standard of care, and no additional follow-up appointments are scheduled strictly for the NAVSaH trial.

### Sample size

Goal enrollment for the pilot NAVSaH trial is 50 patients, based on power calculations to detect significant differences in inflammatory cytokines, radiographic vasospasm, and chronic hydrocephalus.

Under a 2-by-2 repeated measures design consisting of two groups of patients, each measured at two time points, our goal is to compare the change across time in the taVNS group to the change across time in the Sham group. Based upon previous work from Koopman et al. [67], we assume our study will observe 1.1 standardized inflammatory cytokines mean change difference between the two groups. Using a two-sided, two-sample t-test, assuming both time points have equal variance and there is a weak correlation (i.e., 0.15) between measurement pairs, a sample size of 25 in each group achieves at least 80% power to detect a standardized difference of 1.1 in mean changes, with a significance level (alpha) of 0.05 [68].

Based upon our preliminary data, we assume this study will observe 25% and 55% severe vasospasm in the taVNS and Sham groups, respectively. Under a design with 2 repeated measurements (i.e., 2 raters), assuming a compound symmetry covariance structure with a Rho of 0.2, at a significance level (alpha) of 0.05, a sample size of 25 in each group achieves at least 80% power when the null proportion is 0.55, and the alternative proportion is 0.25 [69–71].

As previously described, LV et al. [8] studied the relationship between cytokine levels and clinical endpoints in SAH, including hydrocephalus. From their outcomes, we predict a needed enrollment of approximately 50 to detect these endpoints. From our own preliminary data, with an incidence of chronic hydrocephalus 0% in treated patients and 28.6% in control (despite grade of hemorrhage), alpha = 0.05 and power = 0.80, the projected sample size to capture that change is approximately 44 patients.

### Recruitment

Members of the Neurosurgical team and Intensive Care team identify potential candidates for the NAVSaH trial and notify the research team who conducts formal screening to determine candidacy. Recruitment outside of the hospital, or prior to admission, is not performed.

### Assignment of interventions: allocation

#### Sequence generation

Participants are randomized 1:1 to receive treatment with taVNS or Sham stimulation. Randomization is via a simple randomization via computer generated random generated numbers, with next-number obscured until after consent is completed and time of patient enrollment into the study.

#### Concealment mechanism

The computer-generated randomization sequence is concealed, and the treatment group allocation via random number generation occurs only after the patient has been consented and enrolled in the trial. Following assignment of treatment group, treatment arm remains concealed to the patient, medical team, and outcomes assessors.

#### Implementation

Treatment group allocation is via a simple randomization via computer generated random number assignment (0 vs 1). Patients are enrolled prior to randomization by a member of the research team who goes through the informed consent process with the patient, or their legally authorized representative when appropriate. A single investigator is responsible for entering the patient into the computer system to ascertain the assigned treatment arm.

### Assignment of interventions: Blinding

#### Who will be blinded

Research team members, who apply the ear clips and set stimulation parameters, are not blinded to treatment arm. The participant, the medical team who dictates all management decisions for the patient’s subarachnoid hemorrhage, and outcomes assessors (those who assign modified Rankin scores at admission, discharge, and at each outpatient follow-up appointment) are blinded to treatment arm. Stimulation parameters are sub-sensory, meaning there is no perceived sensation in the setting of stimulation.

#### Procedure for unblinding if needed

There are no proposed circumstances in which unblinding will be performed. If there are concerns regarding adverse events from the intervention then treatment sessions can be ended prematurely, without revealing treatment group.

### Data collection and management

#### Plans for assessment and collection of outcomes

Collected data regarding patient clinical presentation, hospital events, clinical outcomes, results from imaging studies, and serial laboratory data will be collected and stored via REDCap. The web-based interface is designed specifically for clinical trials and prospective observational studies. REDCap and the electronic health record will be used for all data collection.

#### Plans to promote participant retention and complete follow-up

The intervention is only administered during the patient’s hospital stay, so there are no foreseeable barriers to retaining participants during the intervention period. Required blood collection is drawn from an indwelling line when available, or is collected at the same time as a patient’s otherwise collected blood samples if venipuncture is required to minimize additional needle sticks that may dissuade ongoing participation during the hospital stay. All primary endpoints are collected during the hospital stay, with no foreseeable barriers to obtaining this clinical information. Following discharge, the patient is not required to make additional visits. Some exploratory outcomes are tracked following discharged but will not be part of the primary analysis. Not mandating additional study visits minimizes the burden on enrolled patients, and further encourages retention in the study long-term.

#### Data management

Data entry input into RedCap undergoes second user validation to confirm data quality. All analysis of continuous variables includes identification of outliers to assess for errors in data collection or reporting.

#### Confidentiality

During this study, medical history (medical diagnoses, surgical history, current medications), information about a patient’s clinical course during hospitalization (medications or procedures performed, radiology exams obtained, and physical exam findings and functional assessments), as well as laboratory data (samples from blood and cerebrospinal fluid) will be assessed. All of the materials collected are for research purposes only, and data will be kept in strict confidence. No identifiable information will be given to anyone without permission from the subject. The consent form includes the informed consent statement required by Washington University for studies involving PHI. Confidentiality will be ensured by use of unique identification codes. Data will be de-identified and stored with an assigned ID number. Data access will be limited to study staff. Data and records will be kept locked and secured, with any computer data password protected. No reference to any individual participant will appear in reports, presentations, or publications that may arise from the study.

The database will be secured with password protection. The informatics manager will receive only coded information that is entered into the database under those identification numbers. Electronic communication with outside collaborators will involve only unidentifiable information. Additionally, all traceable data from copied medical records will also be removed.

#### Plans for collection, laboratory evaluation and storage of biological specimens for genetic or molecular analysis in this trial/future use

While blood and cerebrospinal fluid samples are collected in this study, there are no current plans for genetic analysis. In this trial, analysis of biological samples is limited to evaluation of inflammatory cytokines as described previously, which are stored labeled only with the deidentified study ID number. However, the informed consent does include an additional request for approval to allow data and collected biological specimens to be used in future research.

## Statistical methods

### Statistical methods for primary and secondary outcomes

The primary outcome of Aim 1 is quantified continuous measures of the plasma and CSF TNF-α collected at two time points, baseline (before treatment) and day 13 after treatment. The taVNS impact on SAH inflammatory markers will be examined via a linear mixed model, where time (i.e., 0- and 13-days post-treatment), treatment (i.e., taVNS vs. Sham), and time-treatment interaction are the fixed effects, and the dependency of measurements clustered within each individual patient will be accounted for via the random effects. Group differences regarding the local and systemic inflammatory cytokines reduction from baseline to day 13 post-treatment will be examined through the interaction of time and treatment effects. Model assumptions will be inspected, and remedy will be applied if necessary. Secondary outcomes involve exploratory analyses of the remainder of the inflammatory markers collected from plasma and CSF.

The primary outcome of Aim 2 is the highest severity of vasospasm (binary, moderate/severe vs. non moderate/severe) as assessed by measurements performed by a Neurointerventionalist. A mixed effect logistic regression model will be used to examine the taVNS and Sham group difference regarding the severity of vasospasm. A generalized estimating equation (GEE) approach will be used to estimate the model parameters assuming a compound symmetry covariance structure, given there might be a possible unknown correlation between observed outcomes within the same patient. For other secondary clinical metrics related to vasospasm, to detect treatment group differences, based upon the distribution of data, independent t-tests or Wilcoxon–Mann–Whitney tests will be performed for continuous variables, and Chi-square tests or Fisher exact tests will be performed for categorical variables.

The primary outcome of Aim 3 is binary (i.e., shunt vs. non-shunt) and the secondary outcome of Aim 3 is continuous (i.e., the duration of external ventricular drainage). To detect the treatment group difference, a 2×2 contingency table and Chi-square test will be used for the binary outcome, and an independent t-test will be used for the continuous outcome.

### Interim analyses

The principal investigators or study staff will review all data collection forms on an ongoing basis for data completeness and accuracy as well as protocol compliance. See Table 1 for data type, frequency of review, and reviewer. A statement reflecting the results of the ongoing data review will also be incorporated into the Annual Report for the Independent Monitor.

The Independent Monitor is appointed to monitor the conduct of the study and subject safety by periodically reviewing data from the study. The Independent Monitor will oversee the overall safety of the study subjects by protecting them from avoidable harm. The Independent Monitor will review adverse events and other relevant study data and will make recommendations regarding continuation of the study to the Principal Investigators. The Principal Investigators will agree to follow the recommendation of the Independent Monitor. This may include termination or suspension of enrollment of the study at any time if it becomes necessary to protect the best interests of the study subjects as advised by the Independent Monitor. If suspicion of an unacceptable risk to participants arises during the conduct of the study, the Independent Monitor, Principal Investigators, institutional review board (IRB), and/or NIH may choose to suspend enrollment until the risk is assessed and a determination is made about the risk. If unacceptable risk is confirmed, the study will be terminated. Decisions to suspend or terminate will be communicated to the IRB. At the time of enrollment suspension or termination a determination will be made about safety follow up assessments required for previously enrolled subjects. The Principal Investigators will notify enrolled subjects, as appropriate, of new information and/or safety follow up assessments as required. Circumstances that may warrant termination or suspension include, but are not limited to: 1) Determination of unexpected, significant, or unacceptable risk to participants, 2) Demonstration of efficacy that would warrant stopping, 3) Insufficient compliance to protocol requirements, 4) Data that are not sufficiently complete and/or evaluable, 5) Determination that the primary endpoint has been met, 6) Determination of futility. The study may resume once concerns about safety, protocol compliance, and data quality are addressed, and satisfy the IRB, Independent Monitor, and NIH.

### Methods for additional analyses (e.g. subgroup analyses)

To evaluate for effect of grade of hemorrhage (high grade with a Hunt and Hess Score ≥3, versus low grade), the interaction between assigned treatment and the subgroup factor in a linear model will be tested (significance level of P<0.017). The model will include a main effect for treatment, subgroup factor, and an interaction between subgroup and treatment. Secondary outcomes will also be assessed in similar models.

### Methods in analysis to handle protocol non-adherence and any statistical methods to handle missing data

High levels of missing data for the outcomes of interest are not expected because these are readily available, documented in medical records as part of the standard of medical care for SAH, and are ascertained during the patient’s primary hospital admission. Should there be high rates of participant attrition in the trial, baseline characteristics and available information on the hospital course will be reviewed and compared to participants who do not withdraw from participation to assess for any differences in those who withdraw and those who do not.

### Plans to give access to the full protocol, participant level-data and statistical code

A dataset with group-level, de-identified clinical data and participant-level de-identified laboratory data in accordance with the Health Insurance Portability and Accountability Act (HIPAA), as well as statistical code used for analyses and will be made available upon reasonable request. The team will comply with the clinical trial information dissemination expectations of the NIH policy and FDA/DHHS Final Rule to register and submit summary results at ClinicalTrials.gov. Consistent with the terms and conditions of NIH funding, we will ensure the submission and updating of registration and results information for this clinical trial in the timeframes established by the Final Rule.

### Oversight and monitoring

#### Composition of the coordinating center and trial steering committee

This is a single-center study. There are no other sites participating in the study and there is no need for a separate administrative or data coordinating center. This center’s study team will recruit and enroll study participants under the delegation and supervision of the site’s Principal Investigators from our Washington University School of Medicine Barnes Jewish Hospital’s patient population.

The research team is comprised of senior investigators in the Departments of Neurosurgery, Biomedical Engineering, and Neurology, who have expertise in the fields of subarachnoid hemorrhage, neuroinflammation, electrical stimulation, and clinical trials. The team’s responsibilities include assessment of trial enrollment progress, overall supervision of the trial, and periodic review of the progress of the study to review safety data.

The lead principal investigator is responsible for overseeing the collection and analysis of data, as well as supervising additional team members who are responsible for identifying potential participants, consenting and enrolling patients, and delivering all treatment sessions.

Communication is facilitated with weekly 1-hour laboratory group meetings where working issues are reviewed, and any difficulties with the technology, data acquisition, transfer, and analysis are addressed. In addition, every week, there is a 2-hour scientific review of the project work. The principal investigators and research team attend these meetings routinely. In addition, an administrative review meeting for budget and grant administration will be undertaken every 3-4 months.

#### Composition of the data monitoring committee, its role and reporting structure

An Independent Monitor who is a Neurosurgical faculty at Washington University who is not associated with this research project and thus works independently of the PI, research team, and funding agencies has been assigned to provide additional periodic review of subject enrollment, clinical endpoints/outcomes, laboratory endpoints, and all reported adverse events. This individual is not a part of the key personnel involved in the study, and is qualified to review the patient safety data generated by this study because of their expertise in the areas of Neurosurgery and Neurology.

#### Adverse event reporting and harms

An adverse event (AE) is any untoward medical occurrence in a subject during participation in the clinical study or with use of the experimental agent being studied. An adverse finding can include a sign, symptom, abnormal assessment (laboratory test value, vital signs, electrocardiogram finding, etc.), or any combination of these.

A serious adverse event (SAE) is any adverse event that results in one or more of the following outcomes:

- Death
- A life-threatening event
- Inpatient hospitalization or prolongation of existing hospitalization
- A persistent or significant disability/incapacity
- A congenital anomaly or birth defect
- An important medical event based upon appropriate medical judgment

SAEs that are unanticipated, serious, and possibly related to the study intervention will be reported to the Independent Monitor and IRB in accordance with requirements.

- Unexpected fatal or life-threatening AEs related to the intervention will be reported to the NIH Program Officer within 7 days. Other serious and unexpected AEs related to the intervention will be reported to the NIH Program Officer within 15 days.
- Anticipated or unrelated SAEs will be handled in a less urgent manner but will be reported to the Independent Monitor, IRB, NIH, and other oversight organizations in accordance with their requirements. In the annual AE summary, the Independent Monitor Report will state that they have reviewed all AE reports.

#### Frequency and plans for auditing trial conduct

**Table 1:**
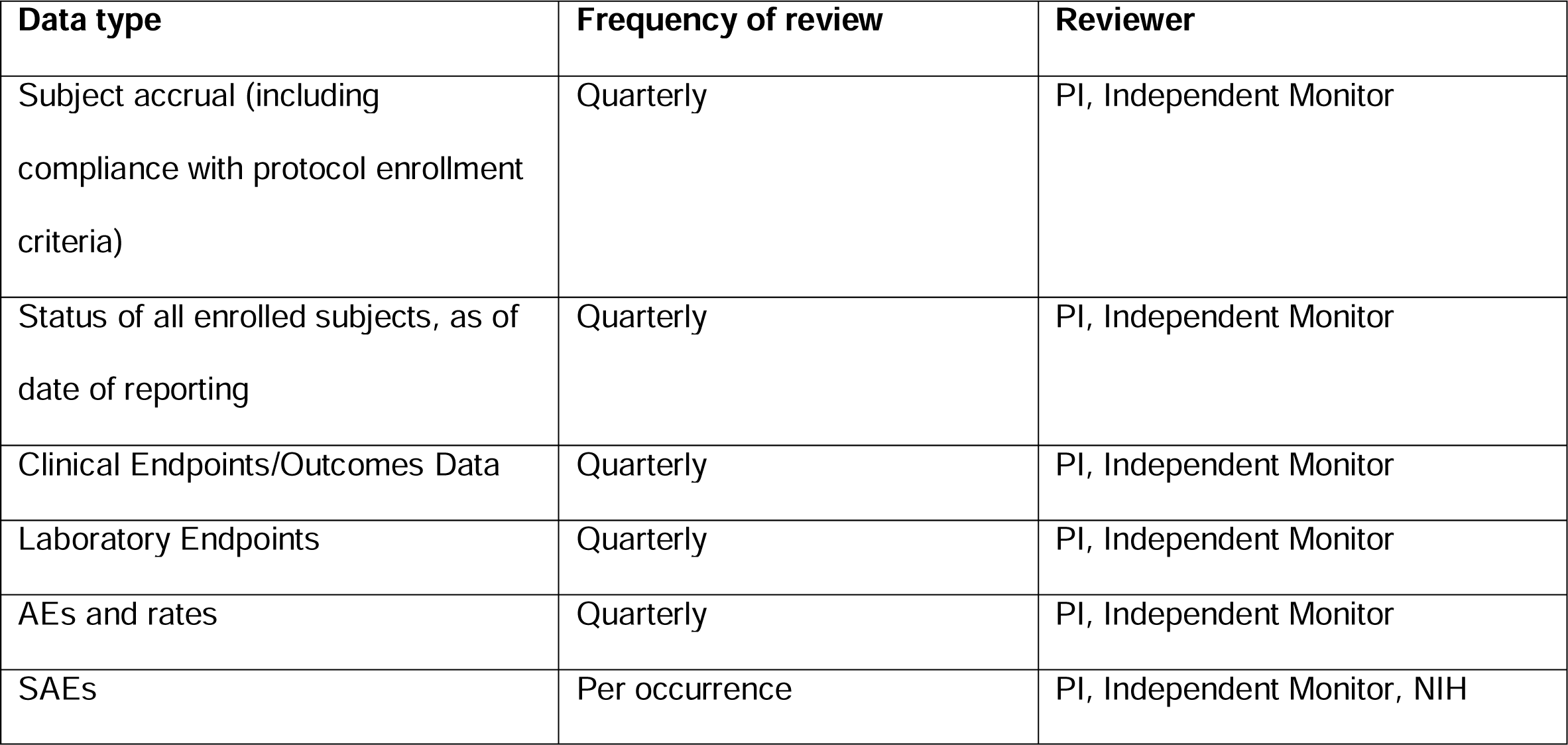
Frequency and personnel for trial conduct auditing.

| Data type | Frequency of review | Reviewer |
| --- | --- | --- |
| Subject accrual (including compliance with protocol enrollment criteria) | Quarterly | PI, Independent Monitor |
| Status of all enrolled subjects, as of date of reporting | Quarterly | PI, Independent Monitor |
| Clinical Endpoints/Outcomes Data | Quarterly | PI, Independent Monitor |
| Laboratory Endpoints | Quarterly | PI, Independent Monitor |
| AEs and rates | Quarterly | PI, Independent Monitor |
| SAEs | Per occurrence | PI, Independent Monitor, NIH |

#### Plans for communicating important protocol amendments to relevant parties (e.g. trial participants, ethical committees)

All protocol amendments will be reviewed by the study investigators, Principal Investigators, and the Washington University IRB committee. This includes main protocol amendments, revisions of any consent paperwork, and continuing reviews. Any protocol amendments that impact enrolled patients will be communicated to patients as deemed appropriate per review and recommendations by the IRB and Independent Monitor.

### Dissemination plans

The Principal Investigators of the clinical trial will comply with the clinical trial information dissemination expectations of the NIH policy and FDA/DHHS Final Rule to register and submit summary results at ClinicalTrials.gov. Consistent with the terms and conditions of NIH funding, we will ensure the submission and updating of registration and results information for this clinical trial in the timeframes established by the Final Rule. Registration and results reporting in ClinicalTrials.gov will be completed within the following timeframes:

- Registration of the trial at ClinicalTrials.gov no later than 21 days after enrolling the first participant.
- All submitted information will be updated at least once a year.
- Any apparent errors, deficiencies, and/or inconsistencies identified by NIH as part of the quality control review process and any other errors identified will be addressed by the responsible party.
- Corrections to submitted information will be made within 15 days for registration information and 25 days for results information.
- Trial results will be submitted no later than one year after the primary completion date.

Informed Consent Documents for the clinical trial will include a specific statement relating to posting of clinical trial information at ClinicalTrials.gov. The Washington University IRB has template language explaining that the study will be posted on ClinicalTrials.gov that investigators are required to include in the consent document for the trial. The Principal Investigators will work closely to ensure that clinical trial registration and reporting occurs in compliance with NIH policies and work together to facilitate the process of registration and results reporting to ClinicalTrials.gov in a timely manner, in keeping within the required timeframes. Once data collection is complete, we will work with our statistician to prepare and submit trial results no later than one year after the primary completion date.

The results of the clinical trial will be disseminated to the public through peer-reviewed publication. Publication will be sought through open access publication to facilitate broad access to the trial’s findings. Authorship in all resulting publications will be assigned according to the International Committee of Medical Journal Editors guidelines

## Discussion

The NAVSaH trial aims to target the deleterious inflammatory response following spontaneous SAH through use of a novel non-invasive neuromodulatory approach with vagus nerve stimulation. In this single-institution, triple-blinded, randomized controlled clinical trial, the investigators aim to evaluate the efficacy of this intervention on inflammatory markers in the blood and cerebrospinal fluid, and on key clinical endpoints including vasospasm, hydrocephalus, and functional outcome. The cumulative result of the proposed project would be both a biological and clinical validation that taVNS can significantly alter SAH-induced central inflammation and its associated clinical comorbidities, with substantial potential to improve the considerable morbidity associated with SAH. This trial will provide a critical foundation of knowledge to build upon, with future studies projected to more deeply understand the mechanism that links ear stimulation to immunomodulation and the stimulation parameters that best optimize the clinical effect.

## Supporting Information

**S1 File** Spirit 2013 Checklist

### Trial status

The clinical trial (protocol v1.12 dated February 09, 2022) began recruitment in December, 2020, and enrollment will take place over 4 years with an estimated enrolment end date of December, 2024, with two year-follow up results completed in December 2026.

### Abbreviations

AE: adverse event
CSF: cerebrospinal fluid
IRB: institutional review board
NAVSaH: Non-invasive Auricular Vagus nerve stimulation for Subarachnoid Hemorrhage
NIH: National Institutes of Health
PI: principal investigator
SAE: serious adverse event
SAH: subarachnoid hemorrhage
taVNS: transcutaneous auricular vagus nerve stimulation
VNS: vagus nerve stimulation

### Authors’ contributions

AH, PB, DL, and EL conceived of, and designed the study. AH, GT, GJ, MA, AC, PB, and EL participated in data acquisition and analysis. AH, GT, MA, TK, JO, AV, DL, GZ, PB and EL participated in interpretation of data. AH, PB, and EL contributed to the drafting and significant revision of the manuscript. All authors approved the submitted version and agree to the accuracy and integrity of this work.

### Funding

This study was supported by the NIH/NINDS under Award Number 1R21NS128307 (to ECL, PB, and GJZ), The American Association of Neurological Surgeons Robert J. Dempsey, MD, Cerebrovascular Resident Research Award (to ALH), The Aneurysm and AVM Foundation Cerebrovascular Research Grant (to ALH), and Just-In-Time (JIT) Core Usage Funding Program from the Washington University Institute of Clinical and Translational Sciences (to ALH and ECL). The funders had no role in the design and conduct of the study, the collection, management, analysis, and interpretation of the data, or the preparation, review, approval of the manuscript, or decision to submit the manuscript for publication.

### Name and contact for trial sponsor

Anna Huguenard, MD

Washington University in St. Louis

### Availability of data and materials

The lead principal investigator and other members of the research team will have full, unrestricted access to and be responsible for the final dataset. Following publication of the primary and secondary results, group-level, de-identified clinical data and participant-level de-identified laboratory data in accordance with the Health Insurance Portability and Accountability Act (HIPAA), as well as statistical code used for analyses and will be made available upon reasonable request.

### Ethics approval and consent to participate

The Washington University Institutional Review Board reviewed and approved the protocol for this trial. In the case of protocol modifications during the trial, approval will be sought from the institutional review board. Written informed consent is obtained from all patients, or their legally authorized guardian in the event the patient is incapacitated from providing their own consent, prior to study enrollment.

### Consent for publication

This manuscript contains no personal identifying information, clinical details, or images that require prior consent for publication. There is no intention of reporting identifying patient information when publishing trial results. Participation information materials and informed consent form are available from the corresponding author upon request.

## Supporting information

Spirit Checklist

IRB approval

IRB approval initial

## Data Availability

All data produced in the present study are available upon reasonable request to the authors

