## Supplementary material for "Non-invasive Auricular Vagus nerve stimulation for Subarachnoid Hemorrhage (NAVSaH): Protocol for a prospective, triple-blinded, randomized controlled trial": Spirit Checklist

| | | Reporting Item | Page Number |
|---|---|---|---|
| **Administrative information** | | | |
| Title | #1 | Descriptive title identifying the study design, population, interventions, and, if applicable, trial acronym | 1 |
| Trial registration | #2a | Trial identifier and registry name. If not yet registered, name of intended registry | 3 |
| Trial registration: data set | #2b | All items from the World Health Organization Trial Registration Data Set | 3 |
| Protocol version | #3 | Date and version identifier | 20 |
| Funding | #4 | Sources and types of financial, material, and other support | 21 |
| Roles and responsibilities: contributorship | #5a | Names, affiliations, and roles of protocol contributors | 20 |
| Roles and responsibilities: sponsor contact information | #5b | Name and contact information for the trial sponsor | 21 |
| Roles and responsibilities: sponsor and funder | #5c | Role of study sponsor and funders, if any, in study design; collection, management, analysis, and interpretation of data; writing of the report; and the decision to submit the report for publication, including whether they will have ultimate authority over any of these activities | 21 |
| Roles and responsibilities: committees | #5d | Composition, roles, and responsibilities of the coordinating centre, steering committee, endpoint adjudication committee, data management team, and other individuals or groups overseeing the trial, if applicable (see Item 21a for data monitoring committee) | 16 |

| | | | |
|---|---|---|---|
| **Introduction** | | | |
| Background and rationale | #6a | Description of research question and justification for undertaking the trial, including summary of relevant studies (published and unpublished) examining benefits and harms for each intervention | 3 |
| Background and rationale: choice of comparators | #6b | Explanation for choice of comparators | 7 |
| Objectives | #7 | Specific objectives or hypotheses | 5 |
| Trial design | #8 | Description of trial design including type of trial (eg, parallel group, crossover, factorial, single group), allocation ratio, and framework (eg, superiority, equivalence, non-inferiority, exploratory) | 6 |
| **Methods: Participants, interventions, and outcomes** | | | |
| Study setting | #9 | Description of study settings (eg, community clinic, academic hospital) and list of countries where data will be collected. Reference to where list of study sites can be obtained | 6 |
| Eligibility criteria | #10 | Inclusion and exclusion criteria for participants. If applicable, eligibility criteria for study centres and individuals who will perform the interventions (eg, surgeons, psychotherapists) | 6 |
| Interventions: description | #11a | Interventions for each group with sufficient detail to allow replication, including how and when they will be administered | 7 |
| Interventions: modifications | #11b | Criteria for discontinuing or modifying allocated interventions for a given trial participant (eg, drug dose change in response to harms, participant request, or improving / worsening disease) | 8 |

| | | | |
|---|---|---|---|
| Interventions: adherance | #11c | Strategies to improve adherence to intervention protocols, and any procedures for monitoring adherence (eg, drug tablet return; laboratory tests) | 8 |
| Interventions: concomitant care | #11d | Relevant concomitant care and interventions that are permitted or prohibited during the trial | 8 |
| Outcomes | #12 | Primary, secondary, and other outcomes, including the specific measurement variable (eg, systolic blood pressure), analysis metric (eg, change from baseline, final value, time to event), method of aggregation (eg, median, proportion), and time point for each outcome. Explanation of the clinical relevance of chosen efficacy and harm outcomes is strongly recommended | 8 |
| Participant timeline | #13 | Time schedule of enrolment, interventions (including any run-ins and washouts), assessments, and visits for participants. A schematic diagram is highly recommended (see Figure) | 10 |
| Sample size | #14 | Estimated number of participants needed to achieve study objectives and how it was determined, including clinical and statistical assumptions supporting any sample size calculations | 10 |
| Recruitment | #15 | Strategies for achieving adequate participant enrolment to reach target sample size | 11 |
| **Methods: Assignment of interventions (for controlled trials)** | | | |
| Allocation: sequence generation | #16a | Method of generating the allocation sequence (eg, computer-generated random numbers), and list of any factors for stratification. To reduce predictability of a random sequence, details of any planned restriction (eg, blocking) should be provided in a separate document that is unavailable to those who enrol participants or assign interventions | 11 |

| | | | |
|---|---|---|---|
| Allocation concealment mechanism | #16b | Mechanism of implementing the allocation sequence (eg, central telephone; sequentially numbered, opaque, sealed envelopes), describing any steps to conceal the sequence until interventions are assigned | 11 |
| Allocation: implementation | #16c | Who will generate the allocation sequence, who will enrol participants, and who will assign participants to interventions | 11 |
| Blinding (masking) | #17a | Who will be blinded after assignment to interventions (eg, trial participants, care providers, outcome assessors, data analysts), and how | 11 |
| Blinding (masking): emergency unblinding | #17b | If blinded, circumstances under which unblinding is permissible, and procedure for revealing a participant's allocated intervention during the trial | 12 |
| **Methods: Data collection, management, and analysis** | | | |
| Data collection plan | #18a | Plans for assessment and collection of outcome, baseline, and other trial data, including any related processes to promote data quality (eg, duplicate measurements, training of assessors) and a description of study instruments (eg, questionnaires, laboratory tests) along with their reliability and validity, if known. Reference to where data collection forms can be found, if not in the protocol | 12 |
| Data collection plan: retention | #18b | Plans to promote participant retention and complete follow-up, including list of any outcome data to be collected for participants who discontinue or deviate from intervention protocols | 12 |
| Data management | #19 | Plans for data entry, coding, security, and storage, including any related processes to promote data quality (eg, double data entry; range checks for data values). Reference to where details of data management procedures can be found, if not in the protocol | 12 |

| | | | |
|---|---|---|---|
| Statistics: outcomes | #20a | Statistical methods for analysing primary and secondary outcomes. Reference to where other details of the statistical analysis plan can be found, if not in the protocol | 13 |
| Statistics: additional analyses | #20b | Methods for any additional analyses (eg, subgroup and adjusted analyses) | 15 |
| Statistics: analysis population and missing data | #20c | Definition of analysis population relating to protocol non-adherence (eg, as randomised analysis), and any statistical methods to handle missing data (eg, multiple imputation) | 15 |
| **Methods: Monitoring** | | | |
| Data monitoring: formal committee | #21a | Composition of data monitoring committee (DMC); summary of its role and reporting structure; statement of whether it is independent from the sponsor and competing interests; and reference to where further details about its charter can be found, if not in the protocol. Alternatively, an explanation of why a DMC is not needed | 17 |
| Data monitoring: interim analysis | #21b | Description of any interim analyses and stopping guidelines, including who will have access to these interim results and make the final decision to terminate the trial | 14 |
| Harms | #22 | Plans for collecting, assessing, reporting, and managing solicited and spontaneously reported adverse events and other unintended effects of trial interventions or trial conduct | 17 |
| Auditing | #23 | Frequency and procedures for auditing trial conduct, if any, and whether the process will be independent from investigators and the sponsor | 18 |
| **Ethics and dissemination** | | | |
| Research ethics approval | #24 | Plans for seeking research ethics committee / institutional review board (REC / IRB) approval | 21 |

| | | | |
|---|---|---|---|
| Protocol amendments | #25 | Plans for communicating important protocol modifications (eg, changes to eligibility criteria, outcomes, analyses) to relevant parties (eg, investigators, REC / IRBs, trial participants, trial registries, journals, regulators) | 18 |
| Consent or assent | #26a | Who will obtain informed consent or assent from potential trial participants or authorised surrogates, and how (see Item 32) | 6 |
| Consent or assent: ancillary studies | #26b | Additional consent provisions for collection and use of participant data and biological specimens in ancillary studies, if applicable | 7 |
| Confidentiality | #27 | How personal information about potential and enrolled participants will be collected, shared, and maintained in order to protect confidentiality before, during, and after the trial | 13 |
| Declaration of interests | #28 | Financial and other competing interests for principal investigators for the overall trial and each study site | 22 |
| Data access | #29 | Statement of who will have access to the final trial dataset, and disclosure of contractual agreements that limit such access for investigators | 21 |
| Ancillary and post trial care | #30 | Provisions, if any, for ancillary and post-trial care, and for compensation to those who suffer harm from trial participation | 8 |
| Dissemination policy: trial results | #31a | Plans for investigators and sponsor to communicate trial results to participants, healthcare professionals, the public, and other relevant groups (eg, via publication, reporting in results databases, or other data sharing arrangements), including any publication restrictions | 18 |
| Dissemination policy: authorship | #31b | Authorship eligibility guidelines and any intended use of professional writers | 20 |
| Dissemination policy: reproducible research | #31c | Plans, if any, for granting public access to the full protocol, participant-level dataset, and statistical code | 16 |

**Appendices**

| | | | |
|---|---|---|---|
| Informed consent materials | #32 | Model consent form and other related documentation given to participants and authorised surrogates | 22 |
| Biological specimens | #33 | Plans for collection, laboratory evaluation, and storage of biological specimens for genetic or molecular analysis in the current trial and for future use in ancillary studies, if applicable | 13 |

Note The SPIRIT Explanation and Elaboration paper is distributed under the terms of the Creative Commons Attribution License CC-BY-NC. This checklist can be completed online using https://www.goodreports.org/, a tool made by the EQUATOR Network in collaboration with Penelope.ai
