## Supplementary material for "Non-invasive Auricular Vagus nerve stimulation for Subarachnoid Hemorrhage (NAVSaH): Protocol for a prospective, triple-blinded, randomized controlled trial": IRB approval

**IRB ID #:** 202007034

**To:** Eric Leuthardt

**Approval Date:** 11/07/23
**Next IRB Approval Due Before:** 04/16/24

**2018 Common Rule/Equivalent Protections** Yes

| **Type of Application:** | **Type of Application Review:** | **Approved for Populations:** |
|---|---|---|
| New Project | Full Board: | Children |
| Continuing Review | Meeting Date: | Signature from one parent |
| Modification | Expedited | Signature from two parents |
| | Exempt | Prisoners |
| | Facilitated | Pregnant Women, Fetuses, Neonates |
| | | Wards of State |
| | | Decisionally Impaired |

Criteria for approval are met per 45 CFR 46.111 and/or 21 CFR 56.111 as applicable.

**Source of Support:**

NIH, National Institute of Neurological Disorders and Stroke (NINDS)
Non-invasive vagus nerve stimulation to mitigate subarachnoid hemorrhage induced inflammation (Eric Leuthardt)
American Association of Neurological Surgeons
Robert J. Dempsey, MD, Cerebrovascular Resident Research Award (Anna Huguenard)
The AVM and Aneurysm Foundation (TAAF)
Cerebrovascular Research Grant (Anna Huguenard)

MATERIALS APPROVED

**Protocol:**

Protocol Version: 6
Protocol Date: 12/14/2020

**Consent/Assent Materials:**

Consent & Assent Forms

Informed consent VNS SAH Rev 11_03_23.rtf

This approval has been electronically signed by IRB Chair or Chair Designee:
Jennifer Kyd, BA, Psychology
11/07/23 1432

**IRB Approval:** IRB approval indicates that this project meets the regulatory requirements for the protection of human subjects. IRB approval does not absolve the principal investigator from complying with other institutional, collegiate, or departmental policies or procedures.

**Recruitment/Consent:** Your IRB application has been approved for recruitment of subjects not to exceed the number indicated on your application form. If you are using written informed consent, the IRB-approved and stamped Informed Consent Document(s) are available in *my*IRB. The original signed Informed Consent Document should be placed in your research files. A copy of the Informed Consent Document should be given to the subject. (A copy of the *signed* Informed Consent Document should be given to the subject if your Consent contains a HIPAA authorization section.)

**Continuing Review:** Federal regulations require that the IRB re-approve research projects at intervals appropriate to the degree of risk, but no less than once per year. This process is called "continuing review." Continuing review for non-exempt research is required to occur as long as the research remains active for long-term follow-up of research subjects, even when the research is permanently closed to enrollment of new subjects and all subjects have completed all research-related interventions and to occur when the remaining research activities are limited to collection of private identifiable information. Your project "expires" at midnight on the date indicated on the preceding page ("Next IRB Approval Due on or Before"). You must obtain your next IRB approval of this project by that expiration date. You are responsible for submitting a Continuing Review application in sufficient time for approval before the expiration date, however you will receive reminder notice prior to the expiration date.

**Modifications:** Any change in this research project or materials must be submitted on a Modification application to the IRB for prior review and approval, except when a change is necessary to eliminate apparent immediate hazards to subjects. The investigator is required to promptly notify the IRB of any changes made without IRB approval to eliminate apparent immediate hazards to subjects using the Modification/Update Form. Modifications requiring the prior review and approval of the IRB include but are not limited to: changing the protocol or study procedures, changing investigators or funding sources, changing the Informed Consent Document, increasing the anticipated total number of subjects from what was originally approved, or adding any new materials (e.g., letters to subjects, ads, questionnaires).

**Unanticipated Problems Involving Risks:** You must promptly report to the IRB any unexpected adverse experience, as defined in the IRB/HRPO policies and procedures, and any other unanticipated problems involving risks to subjects or others. The Reportable Events Form (REF) should be used for reporting to the IRB.

**Audits/Record-Keeping:** Your research records may be audited at any time during or after the implementation of your project. There are Federal, State and Institutional requirements for record retention. Check with your organization and your funding agreement to learn more about what record retention requirements apply to your project.

**Additional Information:** Complete information regarding research involving human subjects is available in the "Washington University Institutional Review Board Policies and Procedures." Research investigators are expected to comply with these policies and procedures and to be familiar with the Belmont Report, 45CFR46, and other applicable regulations prior to conducting the research. This document and other important information is available on the HRPO website http://hrpo.wustl.edu/.
