## Supplementary material for "Non-invasive Auricular Vagus nerve stimulation for Subarachnoid Hemorrhage (NAVSaH): Protocol for a prospective, triple-blinded, randomized controlled trial": IRB approval initial

**Study Title:** Vagus Nerve Stimulation following Subarachnoid Hemorrhage

**Principal Investigator, Co-investigator(s):** Eric Leuthardt, MD

**Sponsor or funding source:** American Association of Neurological Surgeons and The AVM and Aneurysm Foundation (TAAF)

### Background, Rationale and Context

Between 3-5% of adults harbor an intracranial aneurysm, and 18-30% of those individuals have more than one aneurysm[1]. Despite more frequent detection of unruptured aneurysms in the general population, many patients still present initially with a subarachnoid hemorrhage (SAH). For patients presenting with SAH, the mortality rate is 10-25%, with an additional 30% of patients suffering permanent disability[2]. Following SAH, there is significant risk for early brain injury and edema, cerebral vasospasm, and delayed cortical ischemia that all contribute to the high morbidity to these patients.

The pathophysiology of aneurysm formation and rupture is complex, and highly influenced by genetic and environmental factors. Interestingly, there is evidence that systemic and local inflammation may lead to the formation and rupture, as well as poorer outcomes following subarachnoid hemorrhage. T-cell and macrophage-mediated inflammation are believed to mediate some histological changes within the vascular wall that leads to aneurysm formation[3], and macrophage infiltrates in the walls of ruptured aneurysms likely contribute to their fragility[4]. Elevated levels of inflammatory mediators, complement, and vascular cell adhesion molecule-1 (VCAM-1) have also been demonstrated in aneurysms, compared to non-aneurysmal intracranial vessels[5]. In ruptured aneurysms, cathepsin G, a serine protease produced primarily in neutrophils, can be found at the site of aneurysm rupture which again implicated neutrophils in the acute rupture process[4].

Following SAH, blood within the subarachnoid space triggers a local and systemic inflammatory response. Studies have shown that following SAH, there are increases in IL-1b[6], IL-6[7-9], IL-18, and TNF-a[6], within the CSF, increases in IL-1[10], IL-23, IL-17[11], and ICAM-1[10] in the serum, and increases in p-38 and p-MAPK[10] in brain tissue. There is also evidence that inflammatory markers are correlated with patient outcomes. Elevated IL-6 has been associated with increased risk for vasospasm[7] and poorer outcomes[8]. Elevated IL-1b, IL-18, and TNF-α in the CSF are associated with cerebral edema and acute hydrocephalus[6]. There is also evidence that the degree of leukocytosis alone on admission following SAH is associated with worse modified Rankin scores on discharge[2].

An interesting avenue of research has now aimed to better understand, and eventually to target these inflammatory pathways to improve outcomes after SAH[12-15], with numerous anti-inflammatory interventions trialed in humans[15]. In smaller enrollment studies, there has been some promise of outcome improvement with Cyclosporin A[16,17] and various types of steroids (methylprednisolone[18,19], hydrocortisone[20], and dexamethasone[21]). Other medications demonstrated no impact on overall outcomes, like Clazosentan[22], Cilostazol[23], and IL-1 antagonists[24]. In larger trials with >1000 patients Simvastatin[25], Aspirin, non-steroidal anti-inflammatory medications, and thienopyrindines all demonstrated no improvement in outcomes[26,27]. Therefore, a combination of medication side effects and lack of efficacy have prevented many anti-inflammatory medications from broader use in SAH.

Vagal nerve stimulation (VNS) has previously been established to have anti-inflammatory effects, and has been successfully demonstrated in other models of inflammatory conditions like induced neuroinflammation[28], cerebral ischemia/reperfusion[29], rheumatoid arthritis[30], sepsis[31], and inflammatory bowel diseases[32] or colitis[33]. Harnessing its anti-inflammatory effects, VNS has also been used in a mouse model of cerebral aneurysms and SAH. In this study, VNS not only reduced the rupture rate of intracranial aneurysms, but also reduced the grade of hemorrhage if rupture occurred, and improved survival and outcome after SAH[34]. VNS may be applied by cervical neck dissection and placement of a cuff electrode directly on the nerve, or via a transcutaneous approach which has demonstrated good efficacy, with lower morbidity[35,36].

Given these promising results in animal studies of SAH, and the established safety of auricular VNS, the authors propose prospectively studying this intervention in our SAH population. We aim to determine if inflammatory markers in the blood and CSF are impacted in patients treated with VNS, and also track patient outcomes to better understand the impact on morbidity or mortality.

### Objectives

To quantify inflammatory markers in the blood and CSF for patients who present with spontaneous subarachnoid hemorrhage to Barnes Jewish Hospital, and determine if treatment with auricular transcutaneous vagal nerve stimulation alters the levels of inflammatory markers in these patients over time, or impacts their overall outcome.

### Methods and Measures

#### Design

This is a randomized controlled trial to evaluate clinical outcome and laboratory findings in patients who present with spontaneous subarachnoid hemorrhage and are treated with either current accepted management, or accepted management in addition to auricular transcutaneous vagal nerve stimulation.

#### Setting

Patients in this study will be inpatient at Barnes Jewish Hospital

### Data Collection Criteria

Adult patients who present with a spontaneous subarachnoid hemorrhage to Barnes Jewish Hospital will be considered for enrollment in this study. Exclusion criteria will include patients < 18 years old, patients with a presumed traumatic etiology for their SAH, patients undergoing active cancer therapy, or patients with sustained bradycardia on arrival with a heart rate < 50 beats per minute.

#### Sample Size

We anticipate enrolling approximately 80 people into the study, with random assortment to treatment with standard medical care, and standard medical care with auricular vagal nerve stimulation.

### Intervention

Patients enrolled in the trial will be randomized to treatment with or without electrical stimulation via an auricular, transcutaneous vagus nerve stimulator. All patients will be fitted with the device, which will attach via an ear clip to the left ear. Stimulation sessions will occur for 20 minutes twice daily during the inpatient period. Patients assigned to the control arm will have no electrical applied, those treated with stimulation will be treated with the following parameters: frequency 20 Hz, pulse width 250 μm, and varying intensities from 0.4 mA to 2mA (intensity is always titrated to be a level below painful threshold in a patient). The amplitude of stimulation may be reduced if a patient complains of discomfort at the site of stimulation. The site of stimulation will be inspected daily before and after treatment to ensure there is no redness or irritation at the site.

### Outcome Measure(s)

The primary endpoints of the study will be levels of inflammatory markers (IL-1b, IL-2, IL-4, IL-5, IL-6, IL-8, IL-10, IL-12, IL-13, IL-17a, GM-CSF, IFN gamma, TNF-α, white blood cell count) in the blood and CSF (in patients with an external ventricular drain), as well as specific inflammatory cell counts (monocyte count, for example) at time of admission, and every 3 days throughout hospitalization. In a subset of patients, blood (and CSF when an external ventricular drain is in place) will also be collected again following stimulation to look at the more acute changes in these levels. The secondary endpoints will be mortality, radiographic and clinical vasospasm, need for permanent CSF diversion, and clinical outcome assessed via a modified Rankin score. Vasospasm will be assessed by currently implemented protocols, including CT angiogram and catheter angiogram. Data will also be obtained via chart review from outpatient follow up visits to document patient clinical exams and modified Rankin scores. No additional appointments will be made specifically for the research study.

### Analytical Plan

Results will be analyzed initially using descriptive statistics. Comparison between groups will be done using chi square tests for proportions, and t-tests or ANOVA procedures for continuous variables. Regression analysis will be performed to identify independent outcome predictors. Other inferential statistical analysis will be conducted as appropriate.

### Human Subjects Protection

#### Subject Recruitment Methods

Intensive care unit staff as well as physicians in the Department of Neurosurgery will be made aware of this study, and be directed to notify the research group when a patient presents with a spontaneous subarachnoid hemorrhage. The research team will then screen the patients to see if they are eligible for enrollment in the study. Additional patient data, including medical history, laboratory values, and clinical assessments will be obtained from the electronic health records.

#### Informed Consent

Informed consent will be obtained from the patient if they are able, or from the appropriate party that is providing consent for their medical care if they are unable to provide consent. A copy of the paper consent is also submitted.

#### Confidentiality and Privacy

Confidentiality will be protected by collecting only information needed to assess study outcomes, and maintaining all study information in a secure manner. Data will be de-identified and stored with an assigned ID number. Data access will be limited to study staff. Data and records will be kept locked and secured, with any computer data password protected. No reference to any individual participant will appear in reports, presentations, or publications that may arise from the study.

#### Data and Safety Monitoring

The principal investigator will be responsible for the overall monitoring of the data and safety of study participants. The principal investigator will be assisted by other members of the study staff.

#### Reporting of Unanticipated Problems, Adverse Events or Deviations

Any unanticipated problems, serious and unexpected adverse events, deviations or protocol changes will be promptly reported by the principal investigator or designated member of the research team to the IRB and sponsor or appropriate government agency if appropriate.

14. Lucke-Wold BP, Logsdon AF, Manoranjan B, et al. Aneurysmal Subarachnoid Hemorrhage and

Neuroinflammation: A Comprehensive Review. *Int J Mol Sci*. 2016;17(4):497. doi:10.3390/ijms17040497.

26. Nassiri F, Ibrahim GM, Badhiwala JH, et al. A Propensity Score-Matched Study of the Use of

Non-steroidal Anti-inflammatory Agents Following Aneurysmal Subarachnoid Hemorrhage. *Neurocrit Care*. 2016;25(3):351-358. doi:10.1007/s12028-016-0266-6.
